# Living Kidney Donation in Brazil (2010–2023): An Ecological Time-Series of Donor-Recipient Relationship, Waiting List, and Hospital Indicators Compared with Deceased Donation

**DOI:** 10.64898/2026.02.08.26345842

**Authors:** Márcia Bastos Convento, Fernanda Teixeira Borges

**Affiliations:** Federal University of São Paulo (UNIFESP), Paulista School of Medicine, Department of Medicine, Division of Nephrology, São Paulo, SP, Brazil; Cruzeiro do Sul University, Health Sciences, São Paulo, SP, Brazil

**Keywords:** Kidney transplantation, living donors, graft survival, hospital mortality

## Abstract

**Introduction:** Chronic kidney disease imposes a high clinical and economic burden on the Brazilian Unified Health System, and kidney transplantation offers the best prognosis.

**Objective:** To describe trends in living kidney (LD) donation in Brazil (2010–2023), analyzing the donor-recipient relationship and the operational stock-to-annual production ratio on the waiting list, and to compare hospital indicators and estimated patient and graft survival between LD and deceased-donor (DD) kidney transplants.

**Methods:** Descriptive ecological time-series study using aggregated, publicly available data.

**Results:** The waiting list increased by 15% (from 33,253 to 38,258), and the total number of transplants rose by 29% (from 4,656 to 6,047). Data showed an increase in deceased-donor transplants (from 3,001 to 5,189) and a decrease in LD transplants (from 1,655 to 858), with the LD share declining from 35.55% to 14.19% and the per-million-population rate falling from 8.8 to 4.2. Among LD, there was a relative decrease in related donors (from 82.80% to 71.21%), a relative increase in unrelated spouse donors (from 10.57% to 18.65%), and in other unrelated donors (from 6.63% to 10.14%). Comparatively, LD showed better descriptive performance on survival indicators and lower in-hospital mortality, length of stay, and mean Hospital Admission Authorization value.

**Conclusion:** The findings indicate a need for strategies to sustain DD procurement and LD donation.

## 1. Introduction

Kidney diseases affect more than 844 million individuals worldwide and are major contributors to morbidity and premature mortality^1^. Chronic kidney disease (CKD) represents a substantial public health challenge in Brazil and globally^2^. An estimated 6.7% of the Brazilian adult population is affected by CKD, a proportion that nearly triples among individuals aged 60 years or older. Between 2010 and 2023, hospitalizations coded as CKD increased from 84,337 to 140,648, while CKD-related mortality rose from 2.9 to 4.0 deaths per 100,000 inhabitants^2^.

Over the same period, spending by the Brazilian Unified Health System (SUS) on CKD care increased markedly. Outpatient expenditures on hemodialysis and peritoneal dialysis exceeded R$ 38.3 billion between 2010 and 2023, representing an increase of approximately 73% over the analyzed period^2^. Hemodialysis with up to three sessions per week accounted for the largest share of this expenditure, surpassing R$ 27.5 billion^2^. Hospital expenditures also rose substantially, reaching a cumulative total of R$ 3.18 billion between 2010 and 2023^2^.

This pattern is consistent with growing demand and increasing clinical complexity in the care of patients with CKD in Brazil. Population aging, combined with the high prevalence of hypertension, diabetes mellitus, and cardiovascular disease, likely contributes to the consolidation of CKD as one of the leading public health challenges in the contemporary global context^3^. Improvements in care have been associated with longer survival among patients receiving dialysis and with expanded treatment coverage. At the same time, the incorporation of new technologies and shifts in clinical practice require greater planning and resource allocation to effectively organize nephrology care^4^.

In this context, kidney transplantation stands out as a therapeutic modality capable of restoring renal function in patients with advanced CKD and, compared with dialysis, is associated with longer life expectancy, lower morbidity, and better quality of life^5,6^. Among transplant modalities, living-donor (LD) kidney transplantation offers additional advantages over deceased-donor (DD) transplantation, including higher graft survival rates and, in many cases, shorter time to transplantation, making it the preferred option whenever feasible^7,8^. From an organizational perspective, LD donation may help reduce pressure on the waiting list and improve procedural predictability, thereby potentially optimizing resource use and care pathways^9,10^, provided that donor risks are rigorously assessed and monitored.

Despite these well-established benefits, to the best of our knowledge, this is the first study to describe, using a national time series, the evolution of LD donation in Brazil and its composition according to donor–recipient relationship, while simultaneously comparing outcome indicators with those of deceased-donor transplantation. Accordingly, this study aimed to describe trends in kidney transplantation in Brazil from 2010 to 2023, with a particular focus on temporal patterns in LD donation. We examined the LD profile by recipient relationship, trends on the transplant waiting list, and the operational stock-to-annual production indicator. In addition, we compared living- and deceased-donor transplants with respect to estimated patient and graft survival and, at the hospital level, length of stay, in-hospital mortality, and the mean hospital admission authorization (AIH) value.

## 2. Methods

This descriptive, ecological time-series study adopted a quantitative approach and analyzed aggregated, publicly available secondary data from Brazil covering the period from 2010 to 2023. The series was truncated in 2023 because the 2024 report did not provide a detailed breakdown of the LD profile by donor–recipient relationship. Data sources included the Hospital Information System of the Brazilian Unified Health System (SIH/SUS), accessed via DATASUS^11^, the National Transplant System^12^ (SNT), and the Brazilian Transplant Registry of the Brazilian Association of Organ Transplantation^12^ (RBT/ABTO). No individual-level linkage across databases was performed; therefore, comparisons across sources were descriptive in nature and may reflect differences in coverage, operational definitions, and indicator consolidation methods.

From SIH/SUS^11^ (DATASUS), aggregated data were extracted using TABNET by procedure and place of hospitalization for the period 2010–2023 and stratified by macroregion (North, Northeast, Southeast, South, and Central-West). Kidney transplant hospitalizations were identified using SIGTAP codes 0505020092 (kidney transplant from a deceased donor) and 0505020106 (kidney transplant from a living donor) as the main procedure. For each year and macroregion, we obtained the mean hospital length of stay (days), in-hospital mortality rate (%), and the mean Hospital Admission

Authorization (AIH, R$) value for hospitalizations in which kidney transplantation was recorded as the main procedure. To summarize indicators for the 2010–2023 period, AIH-weighted annual means were calculated to reflect yearly procedure volume; as a sensitivity analysis, simple arithmetic means of annual estimates (with equal weight assigned to each year) were also computed. Monetary values correspond to nominal amounts recorded in the AIH and were not adjusted for inflation; therefore, they do not represent real changes in costs over time. As SIH/SUS is an administrative database, changes in procedure or payment tables, billing rules, and recording practices over time may affect comparability across years and macroregions. SIH/SUS captures hospitalizations financed by the SUS based on AIH issuance and processing by accredited or contracted public and private facilities.

In the SNT^12^, the transplant waiting list corresponds to the Technical Registry of active and semi-active potential recipients, as defined in official reports. For this analysis, we used annual totals from the historical kidney transplant waiting list series, which served as the primary metric. The “general waiting list” reported by the SNT was used only for contextual purposes, as applicable, and its composition may vary over time (e.g., inclusion of different organs or tissues in certain reporting periods). These data represent a stock of records in the Technical Registry and do not capture the dynamics of entries to and exits from the waiting list throughout the year.

From the RBT/ABTO^13^, we obtained annual totals of kidney transplants by donor type (living and deceased), the distribution of donor–recipient relationships among LD donations, transplant rates per million population (pmp), and estimates of patient and graft survival following kidney transplantation. The pmp rates were extracted directly from published RBT/ABTO reports, without recalculation. Survival estimates were derived from the RBT/ABTO Survival Registry, initiated in January 2010. They were analyzed at the published follow-up time points (1, 2, 3, 5, 7, 10, and 14 years), following the definitions of follow-up, censoring, and losses, and graft failure described in the registry documentation, including the estimation methods reported by the RBT/ABTO.

To characterize the relationship between waiting-list stock and annual transplant production, we combined, by calendar year, the stock of patients registered in the SNT with the yearly number of kidney transplants performed, as reported by the RBT/ABTO. This approach allowed the construction of operational stock-to-production indicators (e.g., the absolute difference between registry stock and annual transplant volume and derived proportions). These indicators were intended solely for descriptive purposes and were not interpreted as individual probabilities of transplantation, waiting times, or direct measures of unmet demand.

Variables were analyzed descriptively using absolute and relative frequencies and, when applicable, transplant rates per million population, annual percentage change (Δ%), and compound annual growth rate (CAGR). Annual percentage change was calculated as Δ% = [(current-year value − prior-year value) / prior-year value] × 100. The CAGR was calculated using the formula: CAGR (%) = [(final value / initial value)^(1/n) − 1] × 100, where n corresponds to the number of annual intervals in the analyzed period (final year minus initial year). No hypothesis testing was performed, and all results are presented exclusively in descriptive form.

Because this study was based on aggregated, publicly accessible secondary data and did not involve identifiable individual-level information, submission to a Research Ethics Committee was not required, in accordance with National Health Council Resolution No. 510/2016.

## 3. Results

### 3.1. Kidney transplant waiting list: stock, composition, and the stock–flow relationship

Figure 1 illustrates trends in the stock of patients registered in the Technical Registry (transplant waiting list) in Brazil between 2010 and 2023, considering both the total list (solid organs and cornea) and the kidney transplantation subset. Over the study period, both stocks increased: the total waiting list rose from 59,728 to 68,465 patients (+14.6%), while the kidney transplant waiting-list stock increased from 33,253 to 38,258 patients (+15.0%).

**Figure 1.**
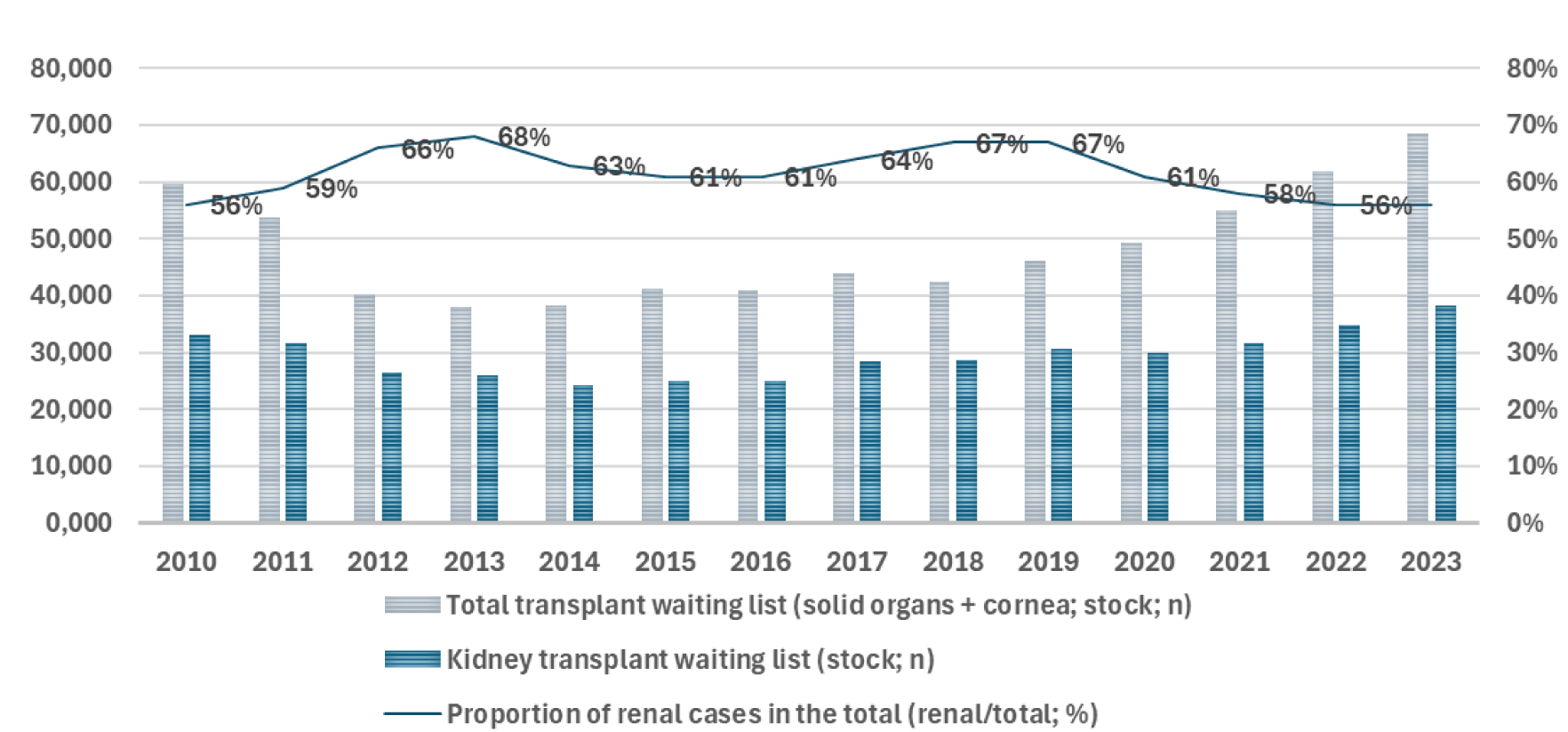
Trends in the stock (Technical Registry) of patients on the total transplant waiting list in Brazil (solid organs + cornea; gray bars; n) and on the kidney transplant waiting list (blue bars; n), and the proportion of kidney patients within the total (line; right axis; %), 2010–2023. Left axis: number of patients (n). Right axis: proportion (%). Note: values represent a stock of registrations and do not capture the dynamics of entries to and exits from the list; they should not be interpreted as an individual probability of transplantation. Data source: SNT [12].

Throughout the series, the proportion of kidney transplant candidates on the total waiting list ranged from 56% to 68%, peaking in 2013 and remaining above 60% from 2012 to 2019. From 2020 onward, this proportion declined through 2022 and remained at 56% in 2023, a value similar to that observed at the beginning of the series. Taken together, these findings indicate that, despite absolute growth in the kidney waiting-list stock, its proportional share of the total waiting list remained relatively stable over time, with a modest decline of approximately 2 percentage points between 2021 and 2023.

These data describe a stock of registrations in the Technical Registry and do not capture the dynamic processes of entries to and exits from the waiting list over time; therefore, they should not be interpreted as reflecting individual probabilities of transplantation.

Figure 2 presents trends in the waiting-list stock (E) of patients listed for kidney transplantation in Brazil (Technical Registry: active and semi-active) and the annual number of kidney transplants performed (T), including both living- and deceased-donor procedures, between 2010 and 2023. Over the study period, the waiting-list stock increased from 33,253 to 38,258 patients (+15.0%), whereas the annual number of transplants performed rose from 4,656 to 6,047 (+29.9%).

**Figure 2.**
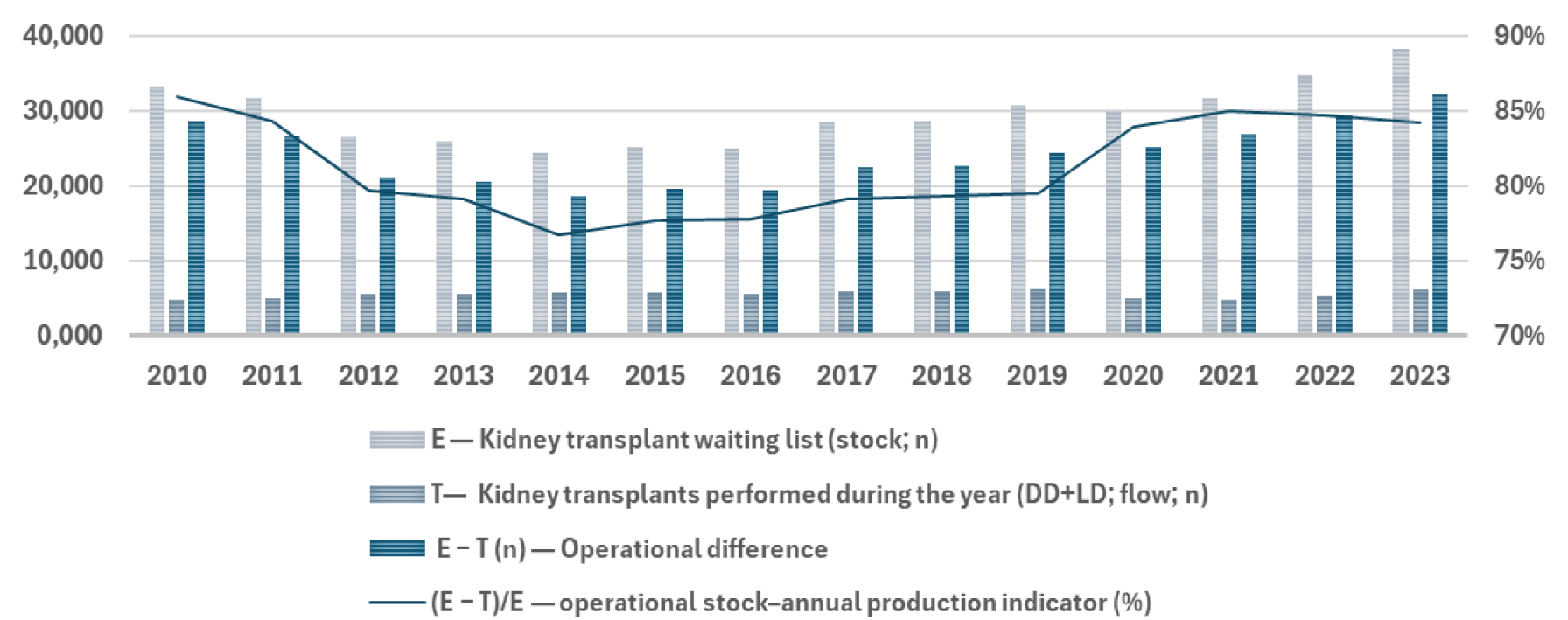
Trends in the waiting-list stock (E) of patients listed for kidney transplantation (Technical Registry: active and semi-active; SNT) and the annual number of kidney transplants performed (T) (living and deceased donors; RBT/ABTO), Brazil, 2010–2023. Bars represent E (n), T (n), and the operational difference E − T (n). The line shows the operational stock–annual production indicator (E − T)/E = 1 − (T/E), in % (right axis; scale expanded for visualization). This is a comparison between the annual recorded stock and the annual transplant production, and it should not be interpreted as an individual probability of transplantation, waiting time, unmet demand, or the “proportion of the list served” in the year. Data sources: SNT [12] and RBT/ABTO [13].

Over the same interval, the operational difference (E − T), defined as the subtraction between the total number of patients on the waiting list reported by the SNT and the number of kidney transplants performed in the year according to the Brazilian Transplant Registry, increased from 28,597 to 32,211 (+12.6%). The operational stock–annual production indicator, expressed as the proportion of the waiting-list stock exceeding the yearly transplant volume, ranged from 77% to 86% across the series. This indicator was 86% in 2010, declined gradually through 2014 (77%), remained relatively stable between 2015 and 2019 (78%–80%), and increased again from 2020 onward (84% in 2020, 85% in 2021 and 2022, and 84% in 2023).

This indicator is operational in nature, as it relates an annual stock measure (waiting-list size) to annual transplant production and does not account for the dynamic processes of entries to and exits from the waiting list within a given year. Therefore, it should not be interpreted as an individual probability of transplantation, an estimate of waiting time, or a direct measure of unmet demand.

### 3.2. Trends in the number of kidney transplants performed with deceased and living donors

Table 1 presents trends in the number of kidney transplants performed with DD and LD in Brazil between 2010 and 2023. Over the study period, the number of DD transplants increased from 3,001 to 5,189 (+72.9%), whereas LD transplants declined from 1,655 to 858 (−48.2%), reflecting opposing trajectories between donation modalities. From 2010 to 2023, the compound annual growth rates were 4.30% for DD and −4.93% for LD.

**Table 1.**
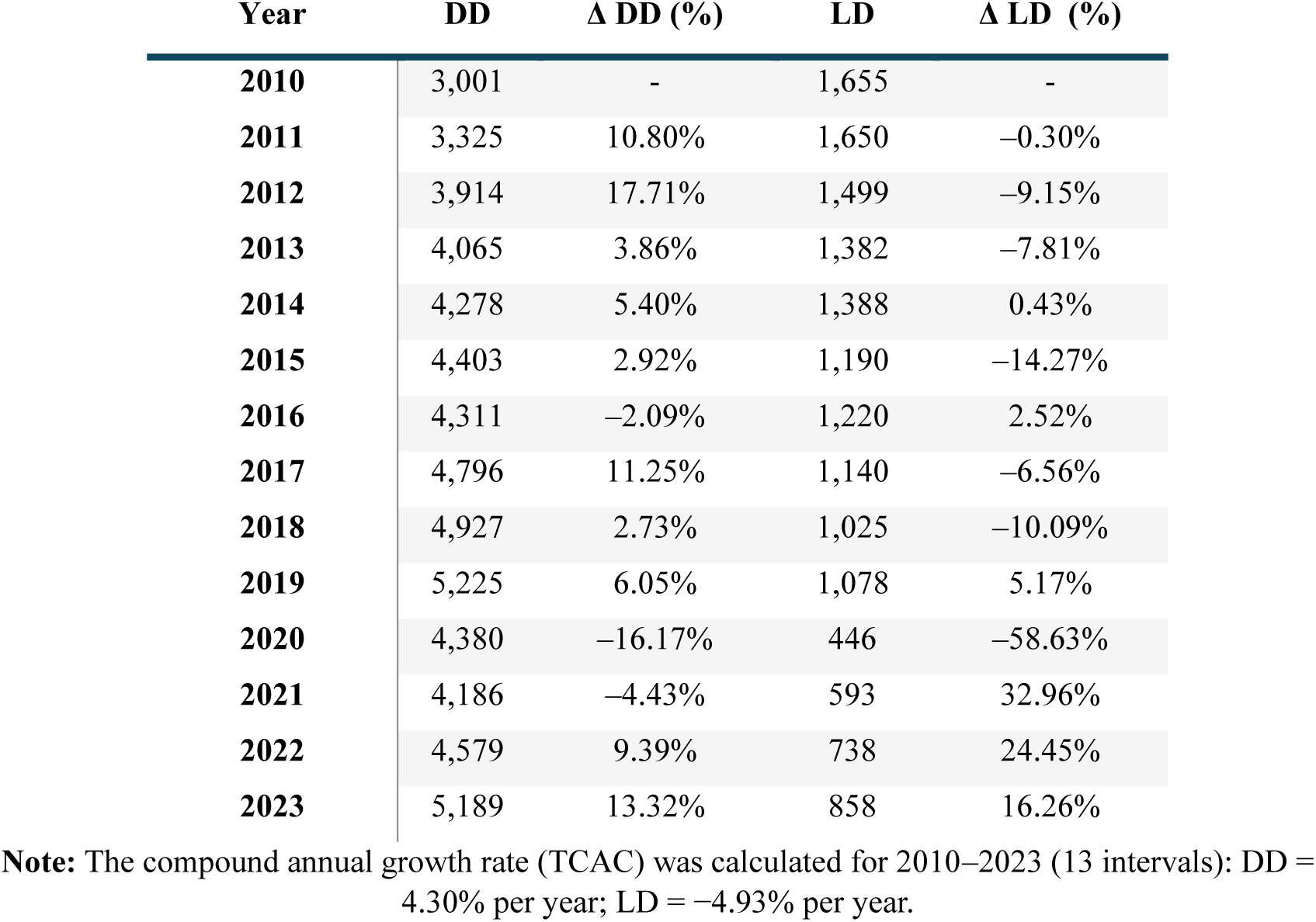
Number of kidney transplants with deceased donors (DD) and living donors (LD) and annual change (Δ%), Brazil, 2010–2023. Data source: RBT/ABTO [13].

Between 2010 and 2015, DD transplants increased steadily, reaching 4,403 procedures, while LD transplants declined to 1,190. In 2016, DD transplants showed a slight decrease (4,311), whereas LD transplants exhibited a one-time increase (1,220). From 2017 to 2019, DD transplants resumed growth, reaching 5,225 procedures, while LD transplants continued an overall downward trend, with minor fluctuations, totaling 1,078 procedures in 2019.

In 2020, the number of kidney transplants declined sharply in the context of the COVID-19 pandemic: DD transplants fell to 4,380 (−16.17% compared with 2019), and LD transplants dropped to 446 (−58.63%). In 2021, DD activity remained below pre-pandemic levels (4,186), while LD began a partial recovery (593 procedures). In 2022, transplant activity increased for both modalities (DD: 4,579; LD: 738). In 2023, DD transplants reached 5,189, the highest value observed in the series, whereas LD transplants increased to 858, remaining below levels observed before 2015.

As a result of these divergent trends, the share of LD transplants among all kidney transplants decreased from 35.6% in 2010 to 14.2% in 2023, indicating a growing predominance of DD kidney transplantation in national transplant activity.

### 3.3. Kidney transplant rates per million population

Table 2 presents trends in kidney transplant rates per million population (pmp) in Brazil by donor type between 2010 and 2023. Over the study period, the overall kidney transplant rate increased from 23.3 to 29.8 pmp, with the highest value observed in 2019 (30.2 pmp). In 2020 and 2021, the overall rate declined to 22.9 and 22.4 pmp, respectively, followed by a subsequent increase to 24.9 pmp in 2022 and 29.8 pmp in 2023. By 2023, the transplant rate had returned to a level comparable to the pre-pandemic period, although it remained below the 2019 peak.

**Table 2.** Kidney transplant rate per million population (pmp), overall and by donor type (DD and LD), Brazil, 2010–2023. Data source: RBT/ABTO [13].

| Year | Kidney transplants<br>(pmp) | DD<br>(pmp) | LD<br>(pmp) |
| --- | --- | --- | --- |
| 2010 | 23.3 | 14.5 | 8.8 |
| 2011 | 24.4 | 16.2 | 8.2 |
| 2012 | 24.5 | 17.3 | 7.2 |
| 2013 | 28.6 | 21.4 | 7.3 |
| 2014 | 29.7 | 22.4 | 7.3 |
| 2015 | 27.6 | 21.7 | 5.9 |
| 2016 | 27.1 | 21.1 | 6.0 |
| 2017 | 28.8 | 23.3 | 5.5 |
| 2018 | 28.7 | 23.7 | 4.9 |
| 2019 | 30.2 | 25.0 | 5.2 |
| 2020 | 22.9 | 20.8 | 2.1 |
| 2021 | 22.4 | 19.7 | 2.7 |
| 2022 | 24.9 | 21.4 | 3.4 |
| 2023 | 29.8 | 25.6 | 4.2 |

**Note:** Rates per million population (pmp) were used as reported in the RBT/ABTO reports.

Rates stratified by donor type followed distinct trajectories. The rate of DD kidney transplants increased from 14.5 pmp in 2010 to 25.6 pmp in 2023. This upward trend was interrupted in 2020 (20.8 pmp) and further in 2021 (19.7 pmp), followed by a recovery in 2022 (21.4 pmp) and the highest rate observed in the series in 2023 (25.6 pmp). In contrast, the rate of LD-kidney transplants declined from 8.8 pmp in 2010 to 4.2 pmp in 2023. The lowest value occurred in 2020 (2.1 pmp), followed by a partial recovery in 2021 (2.7 pmp), 2022 (3.4 pmp), and 2023 (4.2 pmp), remaining below the level observed at the beginning of the series.

Overall, these findings indicate a sustained shift in the kidney donation profile in Brazil, characterized by an increasing relative contribution of DD kidney transplantation over the study period.

### 3.4. Relative share of kidney transplants performed with DD and LD

Table 3 presents the relative share of kidney transplants performed with DD and LD in Brazil between 2010 and 2023. Over the study period, the proportion of transplants performed with DD increased from 64.45% to 85.81%, whereas the share of LD decreased from 35.55% to 14.19%. Consequently, the gap between the two modalities (DD − LD) widened from 28.90 to 71.62 percentage points (p.p.).

**Table 3.**
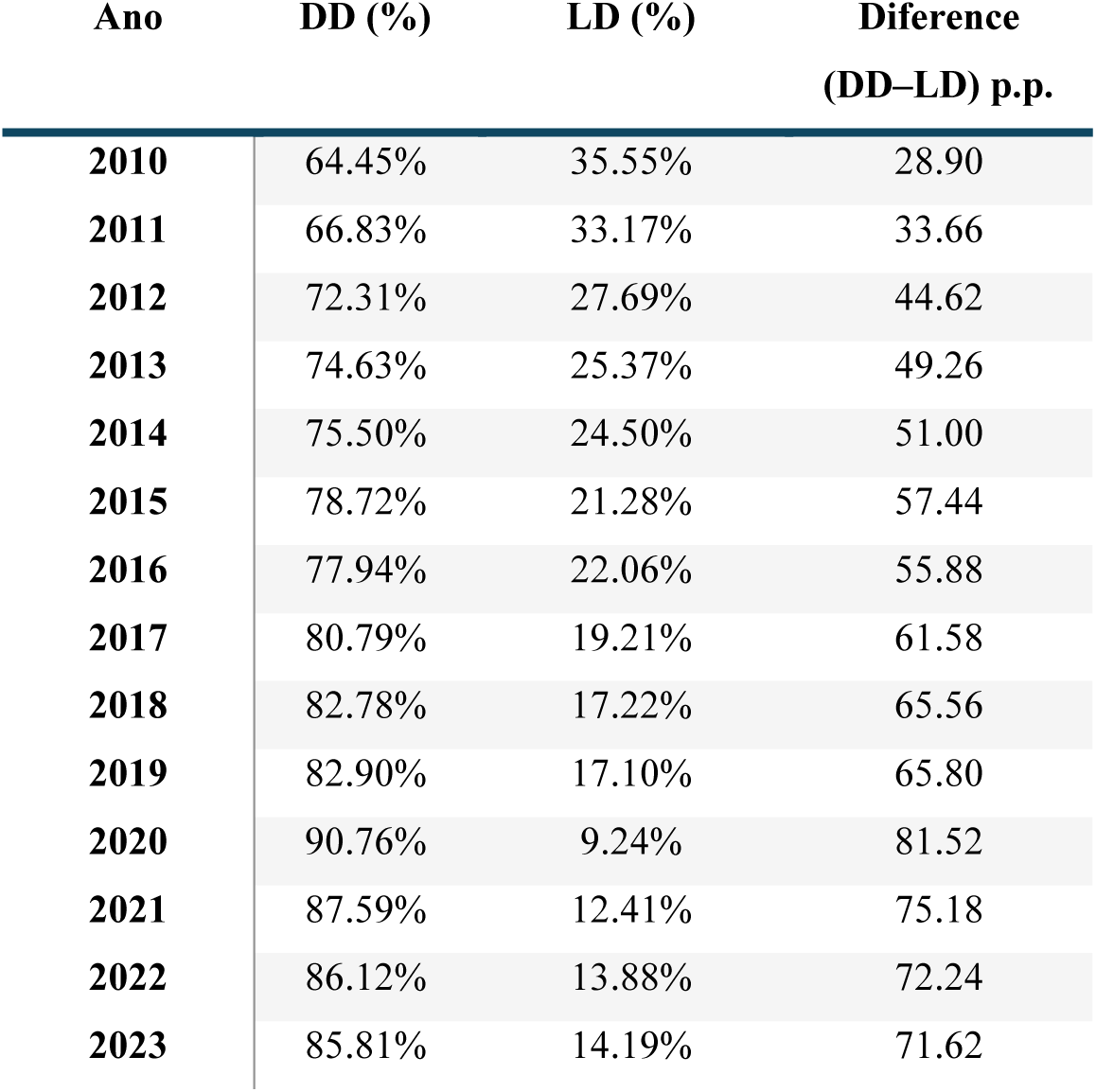
Relative share of kidney transplants with deceased donors (DD) and living donors (LD), and the absolute difference between shares (DD–LD), in percentage points), Brazil, 2010–2023. Data source: RBT/ABTO [13].

| Ano | DD (%) | LD (%) | Diference<br>(DD–LD) p.p. |
| --- | --- | --- | --- |
| 2010 | 64.45% | 35.55% | 28.90 |
| 2011 | 66.83% | 33.17% | 33.66 |
| 2012 | 72.31% | 27.69% | 44.62 |
| 2013 | 74.63% | 25.37% | 49.26 |
| 2014 | 75.50% | 24.50% | 51.00 |
| 2015 | 78.72% | 21.28% | 57.44 |
| 2016 | 77.94% | 22.06% | 55.88 |
| 2017 | 80.79% | 19.21% | 61.58 |
| 2018 | 82.78% | 17.22% | 65.56 |
| 2019 | 82.90% | 17.10% | 65.80 |
| 2020 | 90.76% | 9.24% | 81.52 |
| 2021 | 87.59% | 12.41% | 75.18 |
| 2022 | 86.12% | 13.88% | 72.24 |
| 2023 | 85.81% | 14.19% | 71.62 |

**Note:** Percentages were calculated as $DD/(DD+LD) \times 100$ ; %LD = $100 - \%DD$ ; difference expressed in percentage points (p.p.).

Up to 2015, this gap increased progressively, from 28.90 p.p. in 2010 to 57.44 p.p. in 2015. Between 2016 and 2019, the difference remained high, ranging from 55.88 to 65.80 p.p. In 2020, the gap reached its highest value in the series (81.52 p.p.), coinciding with a sharp decline in LD’s share of total kidney transplants during the COVID-19 pandemic. Between 2021 and 2023, the gap narrowed, reaching 71.62 p.p. in 2023, although it remained above levels observed before 2020.

Overall, these results indicate a sustained increase in the share of kidney transplants performed with DD and a corresponding reduction in the share of LD over the study period.

### 3.5. Profile of living donors by donor–recipient relationship

Table 4 presents trends in LD kidney transplants in Brazil from 2010 to 2023, showing a reduction in overall volume accompanied by a shift in the distribution of donor–recipient relationships. The total number of LD transplants declined from 1,655 in 2010 to 858 in 2023.

**Table 4.** Absolute number (n) and share (%) of living-donor kidney transplants in Brazil, by donor–recipient relationship (related; unrelated-spouse; unrelated-other), 2010–2023. Data source: RBT/ABTO [13].

| Year | Total (LD) | Related (n) | Related (%) | Unrelated-spouse (n) | Unrelated-spouse (%) | Unrelated-other (n) | Unrelated-other (%) |
| --- | --- | --- | --- | --- | --- | --- | --- |
| 2010 | 1,655 | 1,370 | 82.80% | 175 | 10.57% | 110 | 6.64% |
| 2011 | 1,650 | 1,352 | 81.94% | 185 | 11.21% | 113 | 6.85% |
| 2012 | 1,499 | 1,226 | 81.79% | 196 | 13.08% | 77 | 5.14% |
| 2013 | 1,382 | 1,129 | 81.69% | 186 | 13.46% | 67 | 4.85% |
| 2014 | 1,388 | 1,105 | 79.61% | 187 | 13.48% | 96 | 6.92% |
| 2015 | 1,190 | 965 | 81.09% | 166 | 13.95% | 59 | 4.96% |
| 2016 | 1,220 | 963 | 78.93% | 173 | 14.18% | 84 | 6.89% |
| 2017 | 1,140 | 899 | 78.86% | 158 | 13.86% | 83 | 7.28% |
| 2018 | 1,025 | 815 | 79.51% | 148 | 14.44% | 62 | 6.05% |
| 2019 | 1,078 | 833 | 77.27% | 165 | 15.31% | 80 | 7.42% |
| 2020 | 446 | 320 | 71.75% | 82 | 18.39% | 44 | 9.86% |
| 2021 | 593 | 434 | 73.20% | 97 | 16.35% | 62 | 10.45% |
| 2022 | 738 | 535 | 72.50% | 121 | 16.40% | 82 | 11.11% |
| 2023 | 858 | 611 | 71.21% | 160 | 18.65% | 87 | 10.14% |

**Note:** Percentages were calculated relative to the annual total number of living-donor transplants.

Transplants from related donors remained predominant throughout the period. Nevertheless, the absolute number decreased from 1,370 to 611 procedures, and the proportional share declined from 82.80% to 71.21%. Between 2010 and 2015, volumes remained relatively high (from 1,370 to 965), with proportions ranging from approximately 79% to 82%. From 2016 onward, the absolute number declined sharply, reaching 320 in 2020, followed by a partial recovery in subsequent years (434 in 2021 and 611 in 2023). Despite this recovery in volume, the proportional share remained below 74% after 2020, reaching the lowest value in the series in 2023 (71.21%).

Transplants from unrelated spouse donors showed an increase in proportional share over the study period. In absolute terms, this category fluctuated, with a decline in 2020 (82 procedures) followed by a progressive increase, reaching 160 procedures in 2023. Proportionally, the share rose from 10.57% in 2010 to 15.31% in 2019, peaked at 18.39% in 2020, and remained elevated through 2023 (18.65%).

Transplants from unrelated donors classified as “other” declined from 110 procedures in 2010 to 87 in 2023, with the lowest value observed in 2020 (44), followed by a gradual increase in subsequent years. In proportional terms, this category accounted for 6.64% in 2010, ranged from 4.83% to 7.42% between 2012 and 2019, and exceeded 10% from 2021 onward, reaching 10.14% in 2023.

Overall, the results indicate a sustained reduction in the volume of LD transplants, accompanied by an increase in the relative share of donations between unrelated individuals (from approximately 17.2% in 2010 to 28.8% in 2023), particularly among spouses. This proportional shift occurred alongside a decline in the annual LD total, indicating that percentage increases may occur even as absolute numbers decrease.

Figure 3 provides a direct comparison between 2010 and 2023. Over this period, the total number of LD kidney transplants declined from 1,655 to 858, representing an absolute reduction of 797 procedures and a relative decrease of 48.2%. Among related LD, the total fell from 1,370 to 611, corresponding to an absolute reduction of 759 procedures and a decline in relative share from 82.80% to 71.21%. Of the total reduction of 797 procedures, 95.2% occurred among related donors.

**Figure 3.**
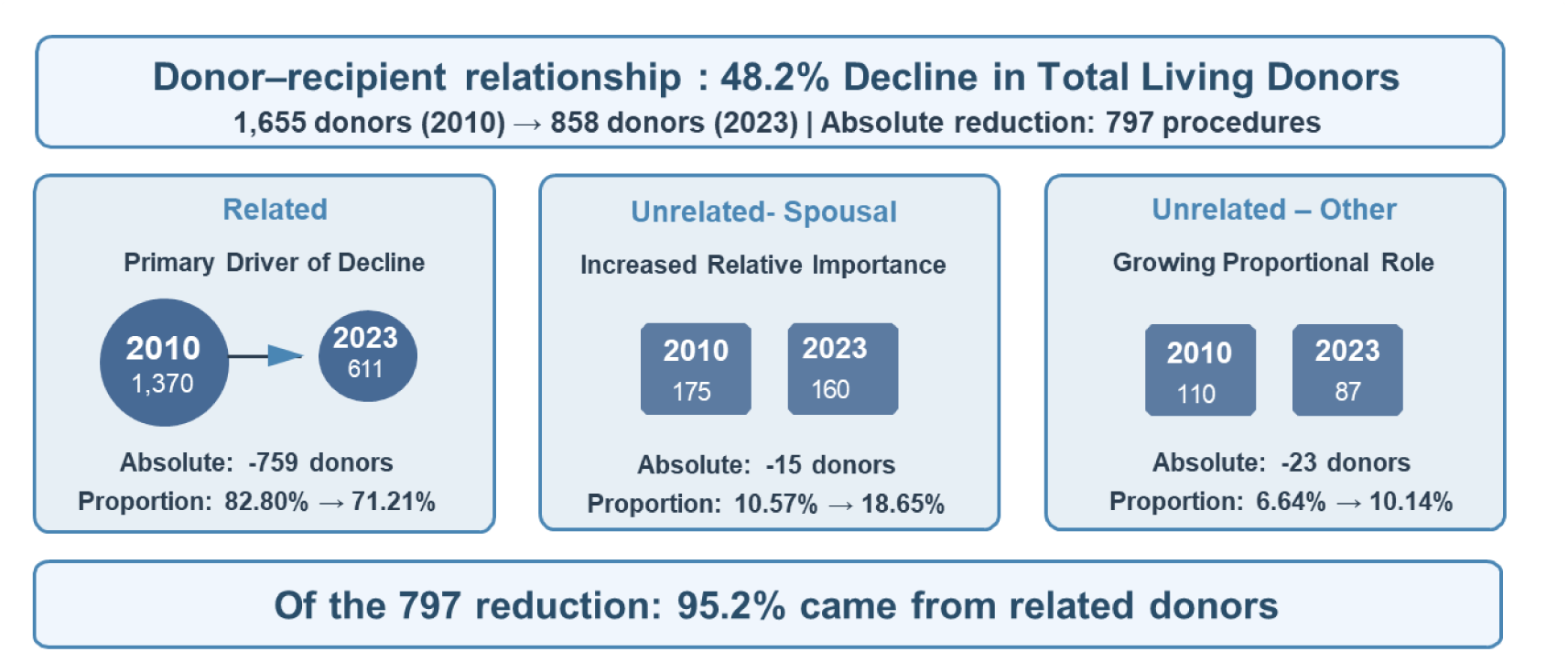
Comparison between 2010 and 2023 of the number of living-donor (LD) kidney transplants in Brazil, by donor–recipient relationship (related; unrelated-spouse; unrelated-other), with absolute (n) and proportional (%) changes. Data source: RBT/ABTO [13].

Among unrelated spouse living donors, the absolute number changed from 175 to 160 procedures, while the relative share increased from 10.57% to 18.65%. For unrelated living donors classified as “other,” the total declined from 110 to 87 procedures (an absolute reduction of 23), while the proportional share increased from 6.64% to 10.14%. Taken together, the reduction in the overall volume of LD transplants was driven primarily by the decline among related donors, whereas spouse and other unrelated donor categories increased their relative participation in living kidney donation despite decreases in absolute numbers.

### 3.6. Estimated Patient and Graft Survival After Kidney Transplantation, by Donor Type

Figure 4 presents estimates of patient and graft survival after kidney transplantation, as reported by the RBT/ABTO, by donor type and at follow-up time points (1, 2, 3, 5, 7, 10, and 14 years). At all evaluated time points, patient survival was higher in the LD group than in the DD group: 97% versus 92% at 1 year (a difference of 5 percentage points); 93% versus 83% at 5 years (10 percentage points); and 83% versus 67% at 14 years (16 percentage points).

**Figure 4.**
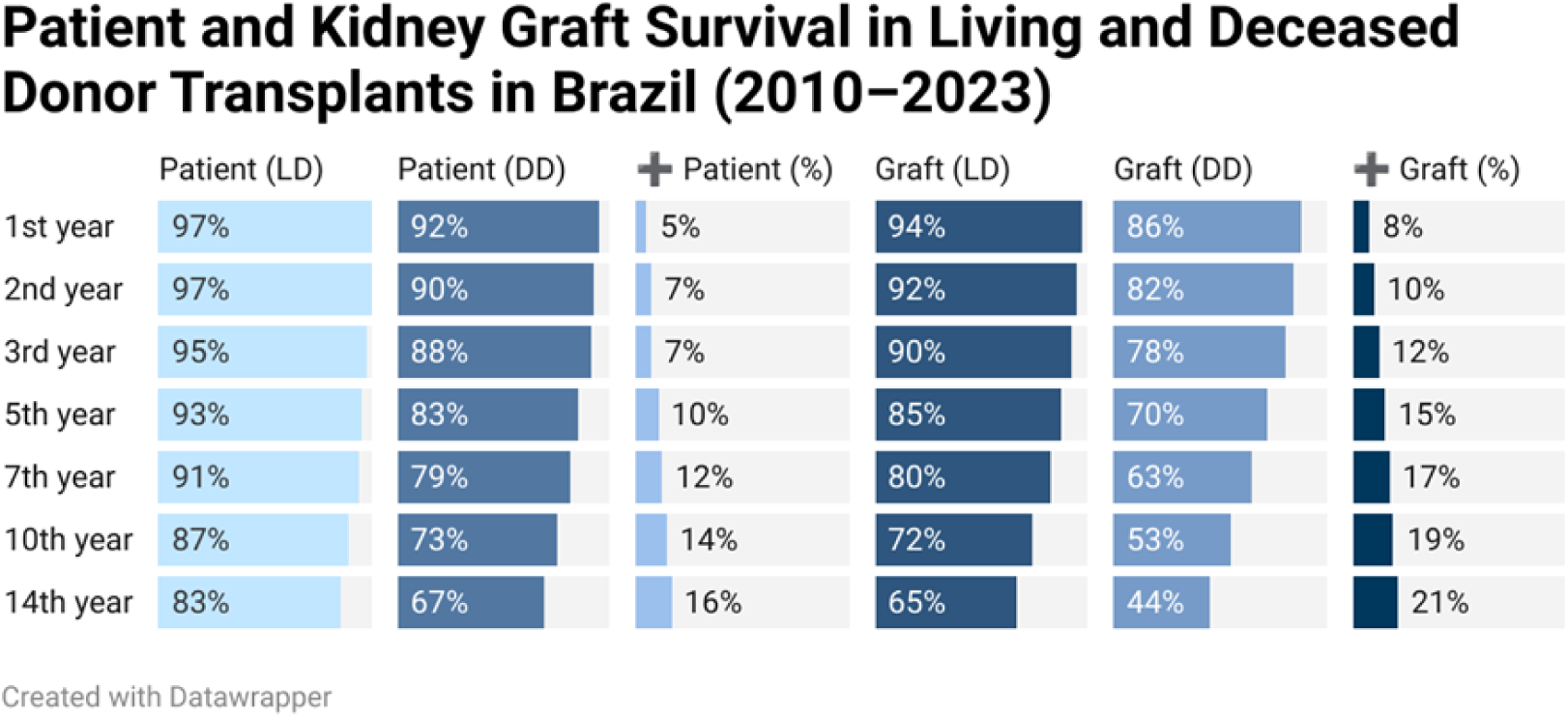
Patient and graft survival after kidney transplantation, by donor type (living donor and deceased donor), at 1, 2, 3, 5, 7, 10, and 14 years of follow-up (values in %), as reported in the RBT/ABTO Survival Registry. Differences between groups are presented in percentage points (p.p.), calculated as LD − DD. Estimates follow the definitions of the follow-up, censoring/loss, and graft-failure criteria described in the RBT/ABTO and are presented descriptively in this analysis. Data source: RBT/ABTO [13].

Graft survival was also consistently higher among LD recipients: 94% versus 86% at 1 year (8 percentage points); 85% versus 70% at 5 years (15 percentage points); and 65% versus 44% at 14 years (21 percentage points). For both patient and graft survival, the differences between donor types widened with increasing follow-up duration.

### 3.7. Hospital indicators by donor type

Table 5 compares hospital indicators by macroregion for SUS-financed kidney transplant hospitalizations, according to donor type, for the period from 2010 to 2023. The indicators include mean length of stay (days), in-hospital mortality (%), and AIH values.

**Table 5.**
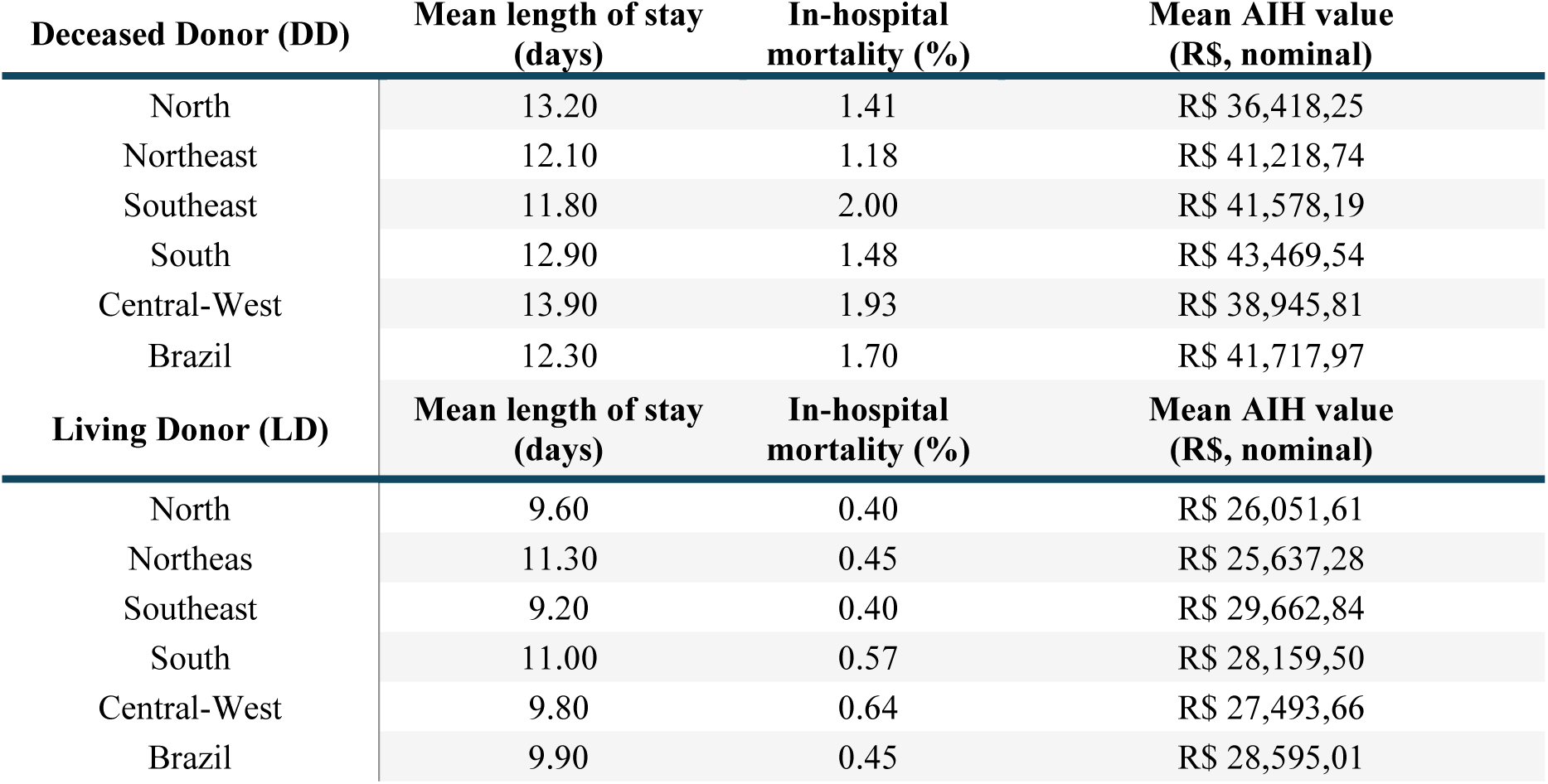
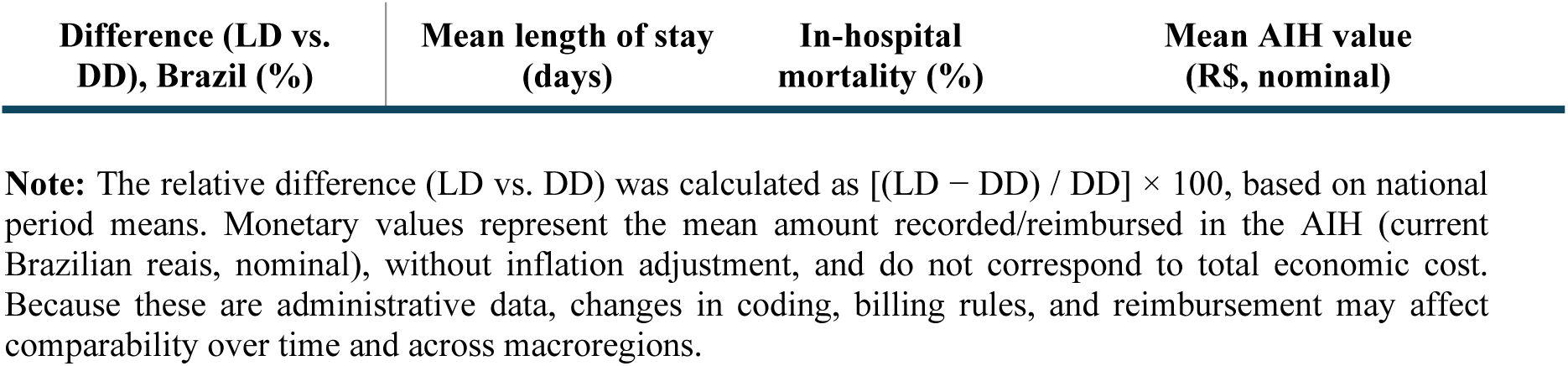
Mean hospital length of stay, in-hospital mortality (%), and mean recorded/reimbursed values (AIH) for SUS-financed kidney transplant hospitalizations, by donor type (DD and LD) and macroregion, Brazil (2010–2023). Data source: SIH/SUS – Ministry of Health [11].

For DD transplants, the mean length of stay ranged from 11.8 days in the Southeast to 13.9 days in the Central-West, with a national mean of 12.3 days. For LD transplants, the mean length of stay was consistently lower across all macroregions, ranging from 9.2 days in the Southeast to 11.3 days in the Northeast, with a national mean of 9.9 days. This corresponds to a relative difference of −19.51% compared with the national mean for DD transplants.

In-hospital mortality for DD transplants ranged from 1.18% in the Northeast to 2.00% in the Southeast, with a national mean of 1.70%. For LD transplants, in-hospital mortality ranged from 0.40% in the North and Southeast to 0.64% in the Central-West, with a national mean of 0.45%, representing a relative difference of −73.53% compared with the national mean for DD transplants.

Regarding mean AIH values recorded and reimbursed, DD transplants ranged from R$ 36,418.25 in the North to R$ 43,469.54 in the South, with a national mean of R$ 41,717.97. For LD transplants, mean AIH values were lower in all macroregions, ranging from R$ 25,637.28 in the Northeast to R$ 29,662.84 in the Southeast, with a national mean of R$ 28,595.01. This corresponds to a relative difference of −31.46% compared with the national mean for DD transplants.

Taken together, these findings describe systematic differences in length of stay, in-hospital mortality, and AIH values between hospitalizations associated with LD and DD kidney transplants. Interpretation of these differences should be undertaken with caution, as they may reflect differences in clinical profiles and selection processes between groups, as well as regional variation in care organization and in SIH/SUS recording and reimbursement practices.

## 4. Discussion

Between 2010 and 2023, the number of patients on the kidney transplant waiting list in Brazil increased by 15%, a magnitude similar to the growth observed in the overall transplant waiting list (14.6%). Although the annual volume of transplant procedures increased over the period, the operational gap between the waiting-list stock and the annual number of transplants also widened, rising from 28,597 to 32,211 patients (+12.6%). At the same time, the proportion of the waiting-list stock exceeding the annual transplant volume remained consistently high (77%–86%). Together, these findings indicate an expansion of transplant activity alongside the persistence of a mismatch between the stock recorded in the Technical Registry and annual transplant production. This represents an operational “stock versus flow” indicator, which does not measure waiting time, individual probability of transplantation, or unmet demand, but is consistent with sustained pressure on the health system.

In this context, describing changes in the composition of organ sources over the series is particularly relevant. Kidney transplant rates per million population (pmp) increased from 23.3 to 29.8 pmp between 2010 and 2023. This growth occurred in parallel with an increase in the rate of DD transplants, from 14.5 to 25.6 pmp, whereas the rate of LD kidney transplants declined from 8.8 to 4.2 pmp over the same period.

This shift is also reflected in the relative distribution of transplant modalities. The share of DD transplants increased from 64.45% in 2010 to 85.81% in 2023, while the share of LD transplants declined from 35.55% to 14.19%. The greatest imbalance was observed in 2020, when the difference between modalities (DD% − LD%) reached approximately 81.5 percentage points (p.p.), coinciding with the sharp reduction in LD participation. In 2023, this difference remained elevated (71.62 p.p.), exceeding pre-2020 levels and indicating a growing predominance of DD kidney transplantation over the historical series.

In the international context, data from the Global Observatory on Donation and Transplantation indicate an approximate global distribution of 62% DD and 38% LD kidney transplants^14^. This pattern contrasts with the Brazilian scenario, in which, in 2023, 85.81% of kidney transplants were performed with DD and only 14.19% with LD, indicating a substantially greater concentration of national transplant activity in postmortem donation compared with the global average.

Understanding this shift involves both population-level and epidemiological factors that may restrict eligibility for LD donation, as well as organizational and regulatory aspects. Between 2010 and 2023, the number of DD transplants increased by 72.9%, whereas LD transplants declined by 48.2%. This pattern is consistent with changes in the epidemiological profile of the adult population, as the rising prevalence of non-communicable chronic diseases increases surgical risk and limits the pool of eligible LD^15^.

National data indicate a high prevalence of excess weight (59.7% of adults), obesity (24.6%), diabetes (10.2%), prediabetes (12%–14%), and hypertension (26.8% of adults, exceeding 50% among older adults) [16–18]. Although the ecological design of this study does not allow causal inference, the temporal concurrence of these trends is consistent with the hypothesis of a progressive restriction in the pool of suitable LD.

In addition, the data reveal changes in donor–recipient relationships among LD. Although related donors remained predominant, their proportional share declined from 82.80% (1,370 transplants) in 2010 to 71.21% (611) in 2023. In contrast, the proportional share of spouses increased from 10.57% (175) to 18.65% (160), and that of other unrelated donors increased from 6.64% (110) to 10.14% (87) over the same period, despite declines in absolute numbers. Descriptively, the reduction in the total volume of LD transplants was concentrated among related donors, resulting in a shift in the relative composition of donor–recipient relationships in a context of declining overall LD activity.

These differences become more relevant when considered alongside care outcomes and performance indicators. Estimates from the RBT/ABTO indicate higher patient and graft survival among LD transplant recipients at all evaluated follow-up time points, with differences widening over time. This pattern is consistent with the literature, but may partly reflect differences in selection processes, clinical profiles, eligibility criteria, and the logistical predictability of elective procedures^16–18^. Accordingly, these comparisons remain descriptive within the scope of this analysis.

In the SIH/SUS database, hospitalizations associated with LD were characterized by shorter lengths of stay, lower in-hospital mortality (0.45% vs. 1.70%), and lower mean AIH (reimbursement) values compared with hospitalizations related to deceased-donor transplantation across all macroregions, in line with international findings^10^. It should be emphasized that AIH values do not necessarily reflect total economic costs and that the figures presented are nominal. Moreover, as these are aggregated and unadjusted estimates, the results should be interpreted as descriptive comparisons that may be influenced by differences in case mix, elective scheduling, logistics, ischemia time, and administrative or recording practices.

Against this background, discussion of the determinants of declining LD and potential mitigation strategies becomes central, with educational interventions emerging as a key component^19–20^. In the Renal Education and Choices at Home (ReACH) study^19^, conducted in the United Kingdom between 2018 and 2020, a pilot home-based education program involving specialist nurse visits to 86 patients, with participation of 141 family members and friends, aimed to increase knowledge, promote dialogue about donation, and reduce barriers to transplantation. Questionnaires administered before and 4–6 weeks after the intervention demonstrated improved knowledge and more favorable attitudes toward transplantation; notably, 53% of patients initiated evaluation of at least one potential donor (78 potential donors).

In a qualitative review of 27 international studies, Truhan et al.^21^ identified late or fragmented provision of information about LD donation as a recurrent barrier, associated with uncertainty, overestimation of donor risk, and difficulties in family communication. The authors emphasized the importance of early, continuous, and professionally mediated education, with early involvement of the family network.

Expansion of LD also depends on regulatory and institutional factors. International evidence highlights the need for multifactorial strategies, including structured public campaigns, integration between dialysis centers and transplant programs, refinement of standards for unrelated donation, and the development of paired-donor programs^22^. Kidney paired donation represents a potential strategy for expanding LD donation in highly structured systems; however, Medina-Pestana et al.^23^ describe barriers to its broad implementation in Latin American and Brazilian contexts, related to socioeconomic inequalities, access asymmetries, and ethical, logistical, and equity challenges. In this context, the authors argue for prioritizing ethically well-established modalities, such as LD donation among consanguineous relatives. Nevertheless, recent initiatives, including Bill No. 3,903/2024, indicate progress in the national regulatory debate on paired donation^24^.

In Brazil, LD donation is regulated by Law No. 9,434/1997 and Decree No. 9,175/2017, which updated and operationalized provisions of the original legislation^25^. For donations between individuals without a kinship relationship, judicial authorization is required to prevent any possibility of organ commercialization. In contrast, donations between consanguineous relatives up to the fourth degree may proceed without judicial authorization^25^. While this legal framework provides important ethical safeguards, it also tends to reduce administrative barriers primarily for consanguineous LD donation, a group that experienced the most pronounced contraction over the study period.

Based on the descriptive findings presented and the contextual evidence from the literature, it is reasonable to consider integrated educational, organizational, and regulatory actions to expand living kidney donation and improve pathways to transplant access, alongside continued strengthening of deceased-donor procurement and strategies to alleviate pressure on the waiting list. These considerations should be interpreted within the ecological and descriptive nature of the analysis, recognizing that the indicators used do not allow for causal inference or direct estimation of individual waiting times or transplantation probabilities.

## 5. Conclusion

Between 2010 and 2023, kidney transplantation in Brazil experienced a shift in donor profile, characterized by an increase in deceased-donor procedures and a reduction in living-donor procedures. In living donation, the data indicate a shift in the composition of donor–recipient relationships, with a more pronounced decline among related donors and an increased relative participation by unrelated donors, particularly spouses.

Taken together, these findings highlight the importance of strategies that both sustain deceased-donor procurement and promote living kidney donation. The descriptive differences observed in survival outcomes (patient and graft) and hospital indicators (length of stay, in-hospital mortality, and mean AIH values) should be interpreted as associations derived from aggregated data, as the study design does not allow causal inference regarding the effect of donor type on these outcomes.

## Study Limitations

This study has limitations inherent to the use of secondary, aggregated, and administrative data, which restrict analyses to the population level and prevent examination of individual determinants of living donation (e.g., age, sex, education, family relationship, and clinical conditions). The use of different information systems (SNT, RBT/ABTO, and SIH/SUS) may lead to divergences in operational definitions, annual closing procedures, completeness, and record consistency, potentially resulting in underreporting, variability across regions and institutions, and information bias. SNT waiting-list measures describe stock in the Technical Registry and do not capture the dynamics of entries and exits over time. In addition, comparisons between donation modalities are descriptive and unadjusted for differences in clinical profile, care complexity (case mix), elective status, and logistical factors. Finally, SIH/SUS indicators reflect hospitalizations within the SUS, with mortality restricted to the hospitalization period, and the mean AIH value represents the recorded/reimbursed amount in nominal terms rather than total economic cost; administrative, coding, and reimbursement changes over the period may affect comparability.

## Authors’ Contributions

Conceptualization: MBC; FTB; Data curation: MBC; Formal analysis: MBC; Investigation: MBC; Methodology: MBC, FTB; Project administration: C, FTB; Resources: C, FTB; Software: MBC; Supervision: FTB; Validation: MBC, FTB; Visualization: MBC, FTB; Writing – original draft: MBC; Writing – review and editing: MBC, FTB. All authors have approved the final version of the manuscript.

## Data Availability

The datasets generated and/or analyzed during the current study are available from the corresponding author upon reasonable request.

## Data Availability

All data referred to in the manuscript are publicly available from official Brazilian health information systems and transplant registries and were accessed in aggregated form.

